# Towards precision well-being in medical education

**DOI:** 10.1101/2023.06.18.23291549

**Authors:** Thomas Thesen, Wesley Marrero, Abigail Konopasky, Matthew Duncan, Karen Blackmon

## Abstract

**Problem:** The escalating mental health crisis among medical students is often met with generalized solutions that overlook substantial individual variations. Furthermore, an exclusive focus on mental illness tends to overshadow the necessity of fostering the positive aspects of medical trainee well-being. This Innovation Report introduces a novel, data-driven precision well-being approach for medical education that is built on a more comprehensive and individualized view of mental health.

**Approach:** Our approach to precision well-being centers on categorizing medical students into distinct and meaningful groups based on their holistic mental health, enabling the future development of tailored wellness support and interventions. We applied k-means clustering, an unsupervised machine learning technique commonly used in precision medicine, to uncover patterns within multidimensional mental health data of medical students. Using data from 3,632 medical students, we formulated our clusters based on recognized metrics for depression, anxiety, and flourishing.

**Outcomes:** Our analysis identified three distinct clusters, each demonstrating unique patterns along the mental health spectrum. Students in the “Healthy Flourishers” cluster expressed no signs of anxiety or depression and simultaneously reported high levels of flourishing, while students in the “Getting By” cluster reported mild anxiety and depression and diminished flourishing. Students in the “At-Risk” cluster expressed high anxiety and depression, minimal flourishing, and increased suicidality. These results represent an integrated, comprehensive empirical model that classifies individual medical students into distinct well-being categories, creating a way for more personalized mental health support strategies.

**Next Steps:** The three-cluster model’s generalizability needs to be improved by incorporating longitudinal data from diverse medical student populations. Integrating physiological markers from wearable devices may improve individualized insights. The model can be used to monitor students’ transitions between clusters, determine influencing factors, form individual risk profiles, and evaluate the effectiveness of personalized intervention strategies stratified by cluster membership.

## Problem

Medical education can engender high levels of psychosocial stress, resulting in elevated rates of anxiety and depression among medical trainees ^1^. High levels of emotional distress can lead to reduction in learning, quality of life and other negative outcomes, including a decline in professionalism and ethics, increased medical errors, relationship and substance abuse issues, and suicide^2,3^. Yet, a deficit orientation towards negative psychological dimensions of medical student mental health may inadvertently overshadow the importance of positive factors, such as well-being, resilience, and flourishing ^4^. A comprehensive understanding of medical students’ mental health requires an exploration of both the positive and negative aspects of mental health, thereby providing a more balanced and holistic perspective that better fits the educational mission.

To create an environment where medical students can thrive and become doctors who can live effective and fulfilling lives, medical educators must take a holistic approach to mental health, considering positive and negative indicators. Yet, no frameworks or empirical models for medical educators exist that unify both ends of the mental health/well-being spectrum. We report here on a novel *Precision Well-Being* framework applied to medical education that addresses this gap. Our proposed framework aligns with the nascent paradigm of *Precision Medical Education*, which has been defined as a “systematic approach that integrates longitudinal data and analytics to drive precise educational interventions which address each individual learner’s needs and goals in a continuous, timely, and iterative fashion, ultimately improving meaningful educational, clinical, or system outcomes” ^5^. Like precision medicine’s objective of tailoring therapeutic strategies to individual patient’s genetic, environmental, and lifestyle factors, precision well-being seeks to understand and address individuals’ mental health—positive and negative—by tailoring support and services to their unique mental health profiles ^6,7^. In this innovation report, we present a machine learning approach to identify clusters of medical students with similar mental health characteristics, preparing the way for more effective and targeted intervention strategies for medical student mental health and flourishing within a precision well-being framework.

## Approach

Our precision well-being innovation involves identifying statistically distinct and conceptually meaningful clusters of medical students based on their self-reported mental health at a discrete time point to route them into personalized wellness interventions. Drawing from the use of unsupervised machine learning in precision medicine, we applied k-means clustering—which groups together data that share certain similarities—as a tool for uncovering hidden patterns and structures in the multidimensional mental health data of medical students. We chose k-means due to its computational efficiency, simplicity of implementation, and ease of interpretation, along with its successful prior application in medical education to understand clerkship grading systems across institutions ^8^. To create our clusters, we used established measures of depression (Patient Health Questionnaire-9 item; PHQ-9), anxiety (Generalized Anxiety Disorder-7 item questionnaire; GAD-7), and flourishing (Diener Flourishing scale, 8 items). The k-means algorithm uses the scores of the individual items from these three measures as input features to group students with similar mental health profiles together, partitioning the data into ‘k’ distinct, non-overlapping groups or clusters. Each student is assigned to the cluster of trainees with the nearest mean (centroid) of mental health scores. This approach allows for integration of both negative and positive mental health aspects into a single empirical model and classification of individual students into distinct categories.

In order to build this model, we used data from the Healthy Minds Study (HMS), which is a comprehensive web-based survey that examines the mental health and related issues of US undergraduate, graduate students, and professional students ^9^. Data from 3,632 students enrolled in US MD programs for the years 2015-2021 were extracted and the responses to the 24 items from the PHQ-9 (9 items), GAD-7 (7 items), and Diener Flourishing Scale (8 items) were used to build the clustering model in Python 3.1. The Healthy Minds Survey was approved by the Health Sciences and Behavioral Sciences Institutional Review Boards at University of Michigan and at all participating campuses, including Dartmouth College. Each scale item was treated as a variable for the analysis, and each individual student’s response constituted a single data point. To determine the optimal number of clusters (k) the Elbow and the Silhouette methods were used based on within-cluster sum of squares and the average silhouette coefficient for each k value, respectively. Paired with a content expert analysis of different cluster solutions, these methods ensured a robust choice of three clusters (k = 3), optimizing the quality of the clusters while minimizing their number. Successful implementation of the clustering model was confirmed through evaluation and labeling of the final clusters by a board-certified psychiatrist. For validation purposes, we performed the k-means cluster analysis on a separate sample of 23,045 students from the HMS data set who were enrolled in a Ph.D. program, which resulted in a highly similar cluster solution (see Appendix).

## Outcomes

Each of the three clusters exhibited unique, explainable patterns along the positive/negative mental health continuum (see Figure 1).

**Figure 1:**
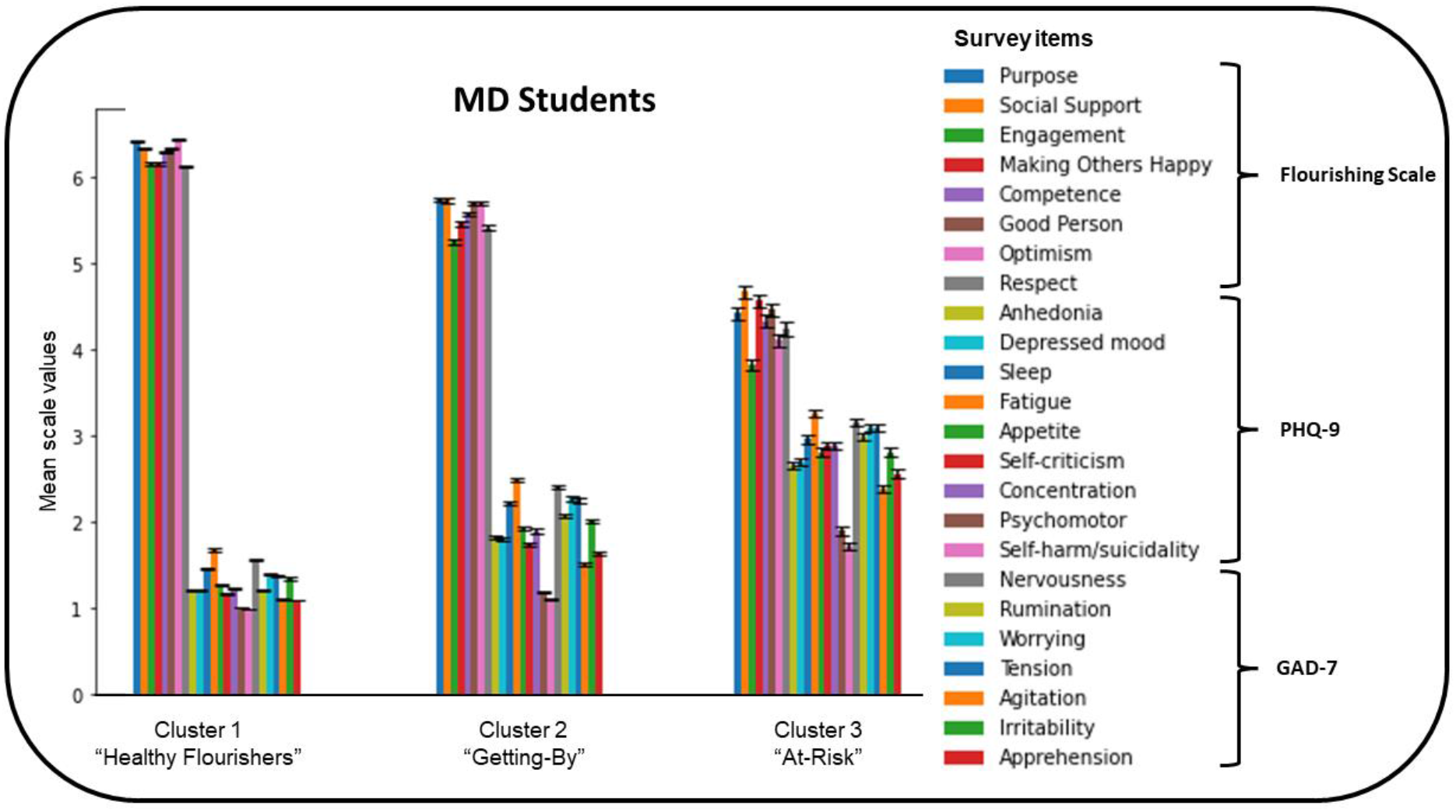
Three cluster solutions with *mean response values from individual survey items for M*.*D. students*. Mean response to each individual item are from the Diener Flourishing Scale (range: 1-7, flourishing), the PHQ-9 scale (range: 1-4, depression), and the GAD-7 (range: 1-4, anxiety) scale. Error bars represent standard error of the mean.

### Cluster 1: Healthy Flourishers

This group is characterized by students who report minimal symptoms of anxiety and depression. They endorse high agreement with statements of self-perceived success in important areas such as relationships, self-esteem, purpose, and optimism (i.e., flourishing). They report slightly more frequent feelings of fatigue (PHQ-9 item #4) and nervousness (GAD-7 item #1) compared to other items. Thoughts of self-harm or suicide (PHQ-9 item #9) were endorsed by 1.3% of students in this cluster.

### Cluster 2: Getting By

This group is characterized by students who report mild anxiety and depression (mainly sleep problems, fatigue, nervousness, worrying, and irritability), and reduced agreement with flourishing statements, relative to cluster 1. Thoughts of self-harm or suicide were endorsed by 11.3% of students in this cluster.

### Cluster 3: At Risk

This group is characterized by students who self-report moderate anxiety, moderate-to-severe depression, and minimal flourishing. Thoughts of self-harm or suicide were endorsed by 48.2% of students in this cluster, with 40% of students reporting thoughts of suicide and self-harm at least half of the time or more frequently.

Within the data set, 49% (n=1780) of medical students were identified by the clustering method as ‘Healthy Flourishers”, 36% (n=1307) as ‘Getting-By’ and 15% (545) as ‘At-Risk’ (see Figure 2).

**Figure 2:**
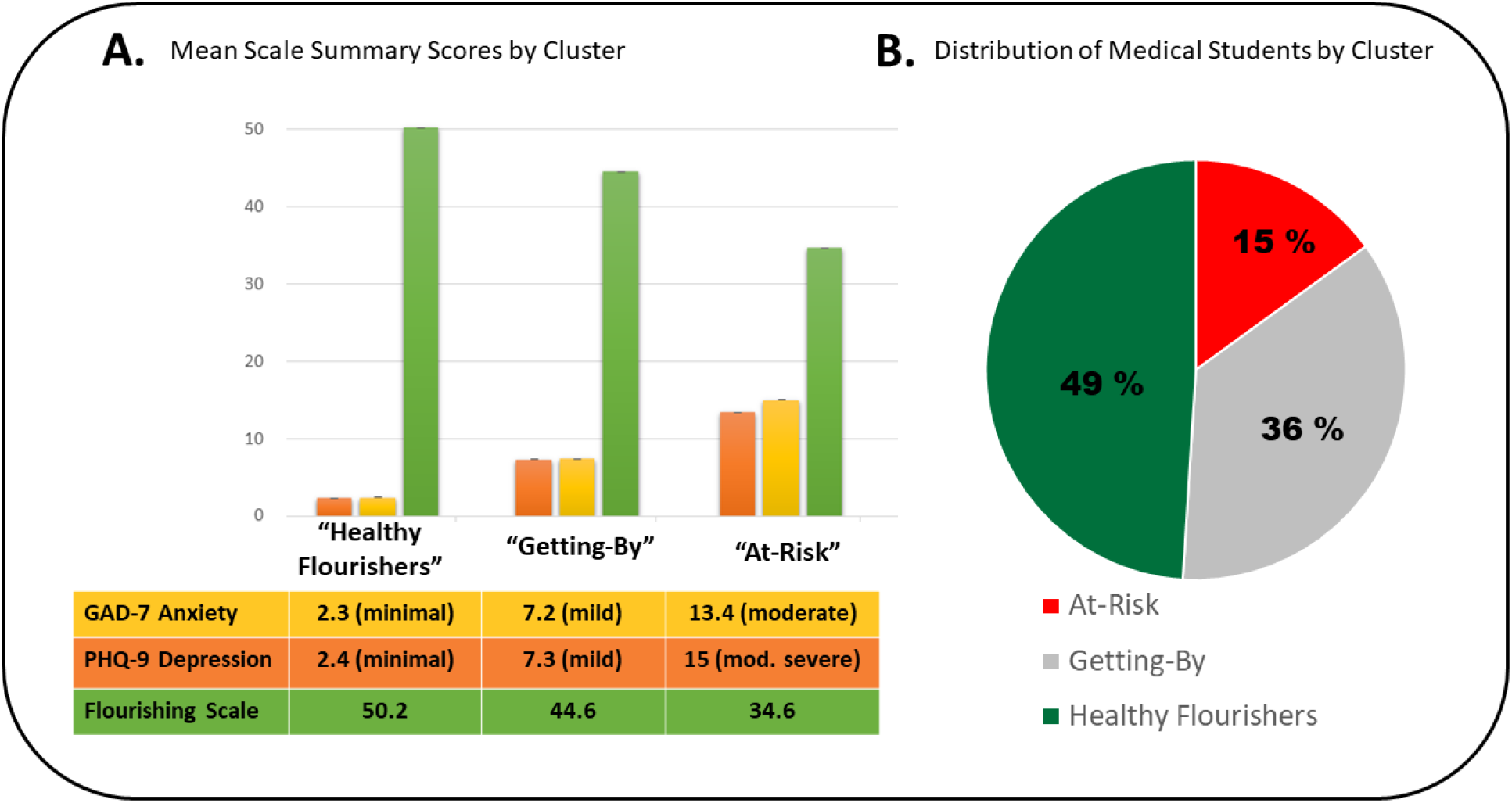
**A**. Mean summary scores and clinical severity labels for GAD-7, PHQ-9, & Flourishing Scale for each cluster of medical students. Error bars represent 95% bootstrapping confidence interval. Note the difference in range for each scale (min-max): GAD-7 from 0-21, PHQ-9 from 0-27, Flourishing Scale from 8-56. For GAD-7 and PHQ-9, higher scores indicate greater symptom reporting. For the Flourishing Scale, higher values indicate greater reported flourishing. **B**. This figure shows the percentage distribution of 3,632 M.D. students among the three clusters. Healthy Flourishers = 1907 students (46%), Getting-By = 1267students (31%), At-Risk = 458 students (11%).

## Implications for mental health support

By applying cluster analysis to a large sample of data collected from diverse US medical schools over a period of six years, we uncovered three unique mental health ‘phenotypes’ and estimated their prevalence among the US medical student population. Notably, almost half of the medical students were classified under the ‘Healthy Flourisher’ category, which is characterized by high flourishing without indicators of frequent psychological distress. These medical students are not merely managing, but indeed are thriving. Such students likely require no interventions other than the recognition and constructive reinforcement of their wellness strategies. Furthermore, they could provide a valuable cohort for research seeking individual social, physiological, and behavioral determinants that foster resilience and flourishing during medical school.

Approximately one-third of medical students were identified within the ‘Getting-By’ cluster, due to endorsing more frequent depressive and anxiety symptoms, which could substantially impact their academic efficacy and holistic well-being. This group of students also endorsed less positive psychological experiences in flourishing domains, such as engagement, sense of competence, optimism, and social support. Although these students may be managing, they are not necessarily maximizing their potential, and could likely benefit from interventions such as coaching or self-guided wellness programs. Around 11% also report suicidal ideation or thoughts of self-harm. It would be essential to monitor this group for any signs of transition into the ‘At-Risk’ category.

The ‘At-Risk’ cluster, constituting around 11 percent of medical students, is distinguished by very frequent feelings of depression and anxiety. Students within this category necessitate referral to counseling services for further evaluation and potential intervention, which could include pharmacotherapy and/or psychotherapy. This group, while the smallest in number, is likely to require the most resources in student and mental health support. Considering the total number of medical school graduations between 2017-2021 of 108,272 ^10^, it can be estimated that in any given year, approximately 16,241 medical students within the US are classified in this vulnerable ‘At-Risk’ category. This finding is important to consider in terms of resource and budget allocation to mental health services within the medical school environment.

The current study is the first to consider the concept of precision well-being in medical education and represents an innovative approach towards defining holistic well-being during medical training. The validity of the resulting clusters is supported by replication of cluster profiles and proportions in an independent sample of Ph.D. students. Implementation of this data-driven model into student wellness programs enables administrators to devise specific interventions tailored to distinct groups of students, based on their well-being profiles. This approach not only enhances the effectiveness of wellness support but also allows for the optimal deployment of resources within Student Affairs, ensuring that efforts are strategically directed towards the students who need them most. The principle of precision well-being as presented here, carries potential for wider applicability, extending to graduate medical education and physician practice. This approach aligns with the principles of precision medicine and holds promise in addressing the escalating concerns surrounding mental health within the medical profession.

## Limitations

Limitations to our analysis include the reliance on voluntary, self-report measures, which may introduce response bias and affect generalizability. Secondly, although our sample is large and spans several medical schools, it may not be representative of all medical students across different regions and racial/ethnic categories, also limiting generalizability. Further data collection and use of fair algorithms are needed to ensure appropriate representation of students currently underrepresented in medicine. Thirdly, our analysis is cross-sectional in nature, collected across all 4 years of medical school, and therefore does not account for potential changes in students’ mental health over time.

## Next Steps

Our future work aims to validate and enhance the three-cluster model by incorporating supplemental data from a variety of medical student demographics, including those currently underrepresented in medicine, over longer time intervals. Additionally, the inclusion of individualized physiological markers from wearable technology, such as physical activity, sleep and heart rate variability could offer further personalized insights, thereby improving the model’s predictive capability and accuracy. Longitudinal studies are required to examine the stability of these clusters and how they may differ across institutions. Studies are also needed to track and predict students’ transitions between clusters over time, providing insights into the personal and systemic factors that influence the evolution of mental health across the curriculum. The refinement of this model should also incorporate application of predictive machine learning algorithms, facilitating the establishment of individualized risk profiles for each student. Ultimately, we hope to conduct assessments of the efficacy of distinct intervention strategies, which will be stratified according to cluster membership, allowing increasingly nuanced, personalized interventions in the future.

## Data Availability

All data produced in the present study are available upon reasonable request to the authors

## Acknowledgements

None

## Ethics approval

The Healthy Minds Survey was approved by the Health Sciences and Behavioral Sciences Institutional Review Boards at University of Michigan and at all participating campuses, including Dartmouth College.

## Disclaimers

None

## Previous presentations

None

