## Supplemental Figures for "Towards precision well-being in medical education"

**Appendix**

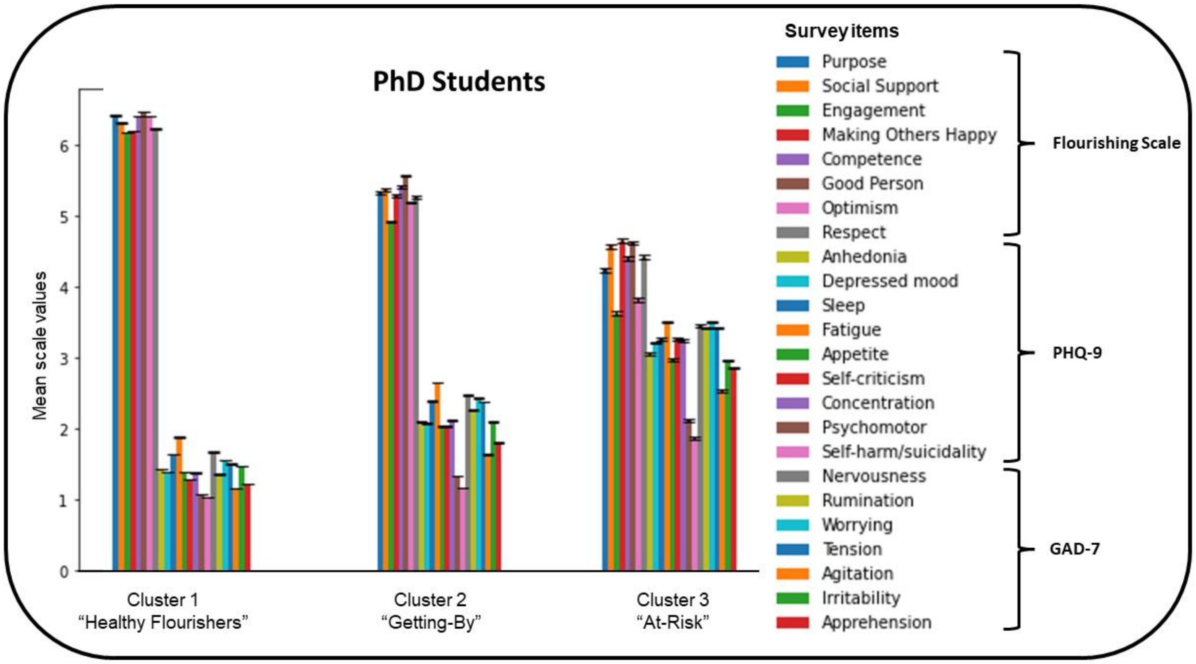


**Figure 2 (appendix):** Mean response values from individual survey items for each cluster for PhD students

This figure shows the mean response from 23,045 PhD students to each individual item from the Diener Flourishing Scale, the PHQ-9 scale, and the GAD-7 scale. Error bars represent standard error of the mean. Cluster properties are highly similar to those generated based on MD student data.

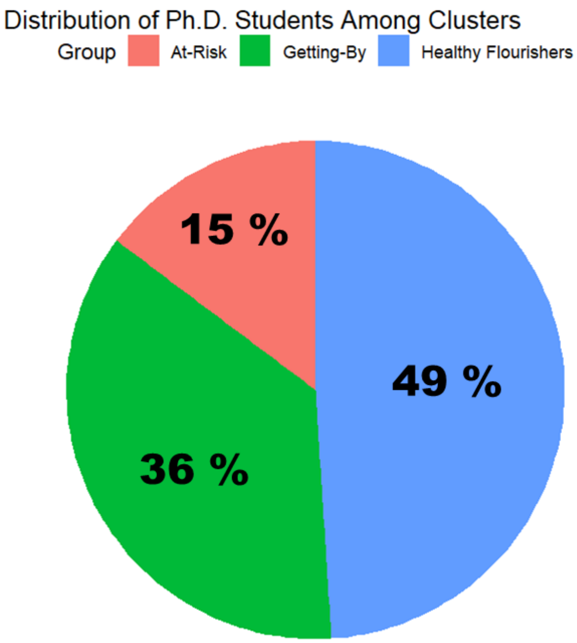


**Figure 2 (Appendix):** Percentage distribution of Ph.D. students among clusters

This figure shows the percentage distribution of 23,045 Ph.D. students among the three clusters, showing a slightly different proportion of students in each cluster compared to the MD students. Healthy Flourishers = 11,288 students (49%), Getting-By = 8,357 students (36%), At-Risk = 3,400 students (15%).
